# Prone positioning of non-intubated patients with COVID-19 - A Systematic Review and Meta-analysis

**DOI:** 10.1101/2020.10.12.20211748

**Authors:** Mallikarjuna Ponnapa Reddy, Ashwin Subramaniam, Zheng Jie Lim, Alexandr Zubarev, Afsana Afroz, Baki Billah, Gabriel Blecher, Ravindranath Tiruvoipati, Kollengode Ramanathan, Suei Nee Wong, Daniel Brodie, Eddy Fan, Kiran Shekar

## Abstract

**Purpose:** Several studies have reported adopting prone positioning (PP) in non-intubated patients with COVID-19-related hypoxaemic respiratory failure. This systematic review and meta-analysis evaluated the impact of PP on oxygenation and clinical outcomes.

**Methods:** We searched PubMed, Embase and COVID-19 living systematic review from December1^st^ 2019 to July23^rd^ 2020. We included studies that reported using PP in hypoxaemic, non-intubated adult COVID-19 patients. Primary outcome measure was the weighted mean difference (MD) in oxygenation parameters (PaO_2_/FiO_2,_ PaO_2_ or SpO_2_) pre and post-PP.

**Results:** Fifteen single arm observational studies reporting PP in 449 patients were included. Substantial heterogeneity was noted in terms of, location within hospital where PP was instituted, respiratory supports, frequency and duration of PP. Significant improvement in oxygenation was reported post-PP: PaO_2_/FiO_2_, (MD 37.6, 95%CI 18.8, 56.5); PaO_2_, (MD 30.4 mmHg, 95%CI 10.9, 49.9); and SpO_2_, (MD 5.8%, 95%CI 3.7, 7.9). Patients with a pre-PP PaO_2_/FiO_2_ ≤150 experienced greater oxygenation improvements compared with those with a pre-PP PaO_2_/FiO_2_ >150 (MD 40.5, 95%CI −3.5, 84.6) vs. 37, 95%CI 17.1, 56.9). Respiratory rate decreased post-PP (MD −2.9, 95%CI −5.4, −0.4). Overall intubation and mortality rates were 21% (90/426) and 26% (101/390) respectively. No major adverse events were reported.

**Conclusions:** Despite significant variability in frequency and duration of PP and respiratory supports, PP was associated with improvements in oxygenation parameters without any reported serious adverse events. Major limitation being lack of control arm and adjustment for confounders. Clinical trials are required to determine the effect of awake PP on patient-centred outcomes.

**Systematic review registration:** Registration/protocol in PROSPERO (CRD42020194080).

**What is the key question?:** Is the novel approach of prone positioning in non-intubated patients associated with improvement in oxygenation?

**What is the bottom line?:** Prone position in non-intubated severe COIVD 19 suffers is associated with improvement of oxygenation while the short- and long-term patient centred out comes in this awake prone patient need more investigation.

**Why read on?:** Our study is first of its kind (Systematic review and Meta-analysis) summarising the evidence surrounding the less invasive innovate technique of prone position in non-intubated COVID-19 patients.

## INTRODUCTION

Coronavirus disease 2019 (COVID-19), caused by the severe acute respiratory syndrome coronavirus 2 (SARS-CoV-2), mainly affects the respiratory system and can lead to acute hypoxaemic respiratory failure. 0.9% to 32% of these patients require admission to intensive care units (ICU) for advanced respiratory support.^1-4^ A surge in critically ill patients with respiratory failure has overwhelmed ICU capacity in many healthcare systems across the world. Studies published during the early phase of the pandemic have showed poor outcomes in invasively ventilated COVID-19 patients. Given a guarded prognosis and significant resource constraints less-invasive, innovative approaches such as prone positioning (PP) of non-intubated patients with hypoxaemic respiratory failure was considered. They were initiated in emergency departments (ED), hospital wards, or in ICUs as an adjunct to conventional oxygen therapies, high-flow nasal cannula (HFNC) and non-invasive ventilation (NIV).^5 6^

The potential efficacy of PP with hypoxaemic respiratory failure is yet to be meaningfully tested in well-designed clinical trials. Limited data suggests that PP in non-intubated patients is feasible and is associated with an improvement in oxygenation in patients with respiratory failure.^7^ There have been case reports and cohort studies that report the use of PP of non-intubated patients with COVID-19 during the pandemic.^2 8-10^ Conceptually, awake PP is relatively less time and resource consuming as compared to PP in intubated patients. Theoretically, they may decrease the risks of adverse events seen in intubated prone patients.

Deteriorating oxygenation despite optimal less-invasive respiratory support^11^ is one of the common triggers for invasive mechanical ventilation. PP improves oxygenation by increasing ventilation– perfusion matching by the recruitment of the larger number of alveolar units located in dorsal areas of the lungs.^12 13^ Furthermore, in patients with COVID-19, PP may also enable gravity assisted diversion of pulmonary blood flows to dorsal regions in the setting of pulmonary vascular dysregulation and loss of hypoxic pulmonary vasoconstriction response in selected patients.^14^ Thus, the success of PP largely hinges on its ability to reliably and predictably improve oxygenation, which may then subsequently improve the respiratory drive, thereby decreasing the risk of self-inflicted lung injury or respiratory fatigue.

Little is known about the magnitude of the effect of PP on oxygenation and its ability to improve patient-centred outcomes in non-intubated COVID-19 patients. Therefore, we performed this systematic review and meta-analysis to evaluate the effect of PP on oxygenation parameters. Secondary analysis included rates of endotracheal intubation and in-hospital mortality.

## METHODS

The protocol for this systematic review and meta-analysis was registered with PROSPERO (CRD42020194080). The study was conducted in adherence with the Preferred Reporting Items for Systematic Reviews and Meta-analyses (PRISMA) Statement.^15^

### Eligibility criteria

Studies on laboratory-confirmed SARS-CoV-2 hypoxaemic adult patients (≥18 years of age) requiring supplemental oxygen who received PP were included. Studies were excluded if (a) they were systematic reviews (b) they did not report on oxygenation parameters (either PaO_2_, SpO_2_ or PaO_2/_ FiO_2_) (c) case reports or case series with fewer than 5 patients (to decrease reporting bias). The corresponding authors of a study were contacted for missing information required for the analysis.

### Search strategy, Information Sources and Study Selection

Two authors (MR and AZ) independently searched on PubMed, Embase, Cochrane, Scopus and the COVID-19 living systematic review from December 1^st^, 2019 to July 23^rd^, 2020. COVID-19 living systematic review has a daily-updated list of pre-print and published articles relating to COVID-19 obtained from PubMed, EMBASE, medRxiv and bioRxiv.^16^ The living systematic review was previously used during the Zika virus epidemic^17^ and recently has been validated against an Ovid search relating to COVID-19 ^18^. Search terms were “Prone”, “Prone Position*” or “Proning” along with “COVID-19”-related terms were used within the title and abstract columns of the systematic review list. Our search was further supported by medical librarian search that was carried out independently (SW). A detailed search terms and tools are summarised in Supplementary Table 1. No language restrictions were applied.

### Quality Assessment and risk of bias in individual studies

The Newcastle-Ottawa Scale (NOS)^19^ was used to assess the quality of cohort studies while Joanna Briggs Institute Critical Appraisal Checklist^20^ was used to evaluate case series. Using relevant appraisal tools, each study was objectively evaluated by two reviewers independently (MR and ZL). Any discrepancies in the approval scores were reviewed and resolved by an additional reviewer (AS) (Supplementary Table 2).

### Study Outcomes

The primary outcome was the change in oxygenation (i.e., PaO_2_/FiO_2_ ratio, PaO_2_ and SpO_2_) following PP. Different variables, such as the saturation of peripheral oxygen (SpO_2_), the partial pressure of arterial oxygen (PaO_2_), and the ratio of PaO_2_ to the fraction of inspired oxygen (PaO_2_/FiO_2_), have been used in the reported studies. We derived the PaO_2_ from SpO_2_ and vice versa if they were not reported in studies using the accepted conversion formulae for consistency to analyse the data (Supplementary Table 3).^21^ For mall number of studies an estimation formula was used to convert median to mean values (Supplementary Table 4).^22^ Median was derived for PaO_2_ in 3 studies, for SPO_2_ in 5 studies and for PaO_2_/FiO_2_ in 2 studies. Sensitivity analyses for physiological parameters were performed by restricting to studies with sample sizes ≥20.

The secondary outcomes included endotracheal intubation rate and mortality. Major adverse events were defined as cardiac arrest, clinically significant haemodynamic instability or accidental dislodgment of intravenous line following PP. Further subgroup analyses were performed to compare: (1) the primary outcome between patients with pre-PP PaO_2_/FiO_2_ >150 and PaO_2_/FiO_2_ ≤150; and (2) the primary and secondary outcomes in patients depending on the location within hospital where PP was initiated (within ICU vs. outside ICU). We also performed an exploratory analysis on the changes in patients’ respiratory rate (RR) after PP.

### Data Analysis

Statistical analyses were performed using the statistical software package Stata-Version 16 (Statacorp, USA). Mean (standard deviation [SD]) or median (interquartile range [IQR]) were used for numerical data and proportion for categorical data. We report weighted mean difference (MD) with 95% confidence intervals (95%-CI) for physiological parameters and event rates using a random effects model to account for both within-study and between-study variances.^23^ Results were presented in Forest plots. Heterogeneity was tested using the χ^2^ test on Cochran’s Q statistic, which was calculated using H and I^2^ indices. The I^2^ index estimates the percentage of total variation across studies based on true between-study differences rather than on chance. Conventionally, I^2^ values of 0–25% indicate low heterogeneity, 26–75% indicate moderate heterogeneity, and 76–100% indicate substantial heterogeneity.^24^ A subgroup analysis using different sample sizes was carried out to identify the possible causes of substantial heterogeneity.^24^ Due to concerns of the limited available data we could not pre-specify the exact variables for subgroup analysis. Following data collection, we carried out two subgroup analyses on oxygenation and clinical outcomes-: ICU vs. non-ICU (emergency department [ED], respiratory wards, high dependency units [HDU]) and baseline PaO_2_/FiO_2_ ratio (PaO_2_/FiO_2_ ≤150 and >150). Symmetry of the funnel plots was evaluated, and the Egger’s regression test was used to examine for publication bias.^25^ A p-value <0.05 was considered significant.

## RESULTS

From 248 studies we identified 15 eligible studies^2 9 10 26-37^ and a total of 449 patients were included in the final analysis (Figure 1) The 15 included studies are summarised in Table 1. The reports originated from 6 countries (China, France, Iran, Italy, USA and UK). 287 patients were men (63.9%) with a mean age (SD) of 56 (7) years. The patients received PP for a variable duration (median 180 minutes, IQR 37.5-264.75) and this procedure was repeated 1-13 times/day during their hospital stay or until intubation, if it occurred. Data on oxygen therapy provided during PP was reported in 350 patients. 68.9% (241/350) received NIV, 4.9% (17/350) on HFNC, 13.7% (48/350) received oxygen via face mask, 12.6% (44/350) via low-flow nasal cannula. Among the 277 patients for whom FiO_2_ was reported, 175 (63.2%) of them received FiO_2_ <50%, 46 (16.6%) were on FiO_2_ between 50-70% and 56 (20.2%) of them received FiO_2_ >70% (Supplementary Table 5).

**Figure 1:**
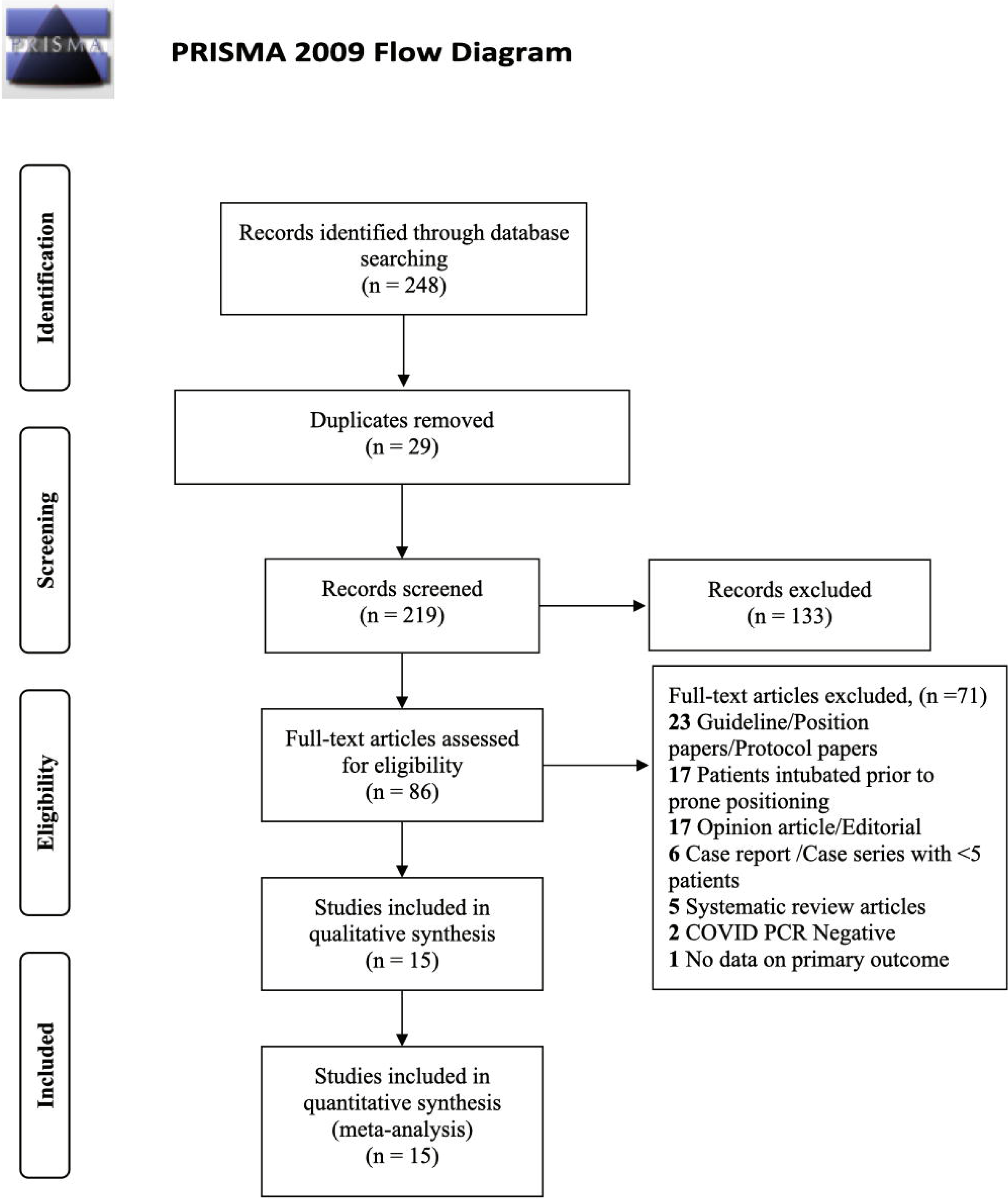
PRISMA flowchart of study inclusions and exclusions. *From:* Moher D, Liberati A, Tetzlaff J, Altman DG, The PRISMA Group (2009). *P*referred *R*eporting *I*tems for *S*ystematic Reviews and *M*eta-*A*nalyses: The PRISMA Statement. PLoS Med 6(7): e1000097. doi:10.1371/journal.pmed1000097 **For more information, visit www.prisma-statement.org**.

**Table 1.** Studies included in the systematic review and meta-analysis.

| Author,<br>reference | n* | Settings | Patient<br>location of<br>PP | Supplemental<br>oxygen and<br>non-invasive<br>respiratory<br>support | Number of<br>episodes,<br>And<br>duration of<br>PP (hours) | Mean<br>duration of<br>PP when<br>respiratory<br>parameters<br>were assessed<br>(minutes) | Respiratory physiology<br>parameters reported<br>pre- and post PP |  |  | Other outcome parameters reported |  |  |
| --- | --- | --- | --- | --- | --- | --- | --- | --- | --- | --- | --- | --- |
|  |  |  |  |  |  |  | P/F<br>ratio | RR | SpO<sub>2</sub> | Hospital<br>mortality | Patients<br>requiring<br>intubation | Hospital<br>length of<br>stay |
| Caputo et al <sup>2</sup> | 50 | Single Centre,<br>NY, USA. | ED | NRB and NC | 1, (NR) | 5 | D | NR | + | NR | + | NR |
| Coppo et al <sup>26</sup> | 46 | Single Centre,<br>Monza, Italy | ED,<br>Respiratory<br>HDU | NIV, VM and<br>NRB | 1-3, (3.5 hrs) | 10 | + | + | + | + | + | NR |
| Damarla et al <sup>27</sup> | 10 | Single Centre,<br>Baltimore, USA | ICU^ | HFNC and NC | Multiple (2<br>hrs) | 60 | D | + | + | 0 | + | NR |
| Despres et al <sup>10</sup> | 6 | Single Centre,<br>Besancon, France | ICU | HFNC or VM | Multiple (1-7 hrs) | 180 | + | NR | D | NR | + | NR |
| Dong et al <sup>37</sup> | 25 | Single Centre,<br>Wuhan, China | ICU | HFNC, VM,<br>NC and NIV | Daily (4.9) | 294 | + | + | NR | 0 | 0 | NR |
| Elharrar et al <sup>36</sup> | 24 | Single Centre,<br>France | NR | NC and HFNC | <1h, 1-3hrs,<br>>3hrs | 90 | D | + | D | NR | + | NR |
| Golestani-Eraghi et al <sup>35</sup> | 10 | Single Centre,<br>Teheran, Iran | ICU | NIV | NR/multiple<br>(14 hr) | NR | + | NR | D | + | + | NR |
| Lawton et al <sup>28</sup> | 165 | Single Centre<br>Bradford, UK | Ward, ED | NIV | 2 times/day | 30 | + | + | + | + | + | NR |
| Moghadam et al<br><sup>34</sup> | 10 | Single Centre,<br>Qom, Iran | ICU | NR | NR | NR | NR | + | + | NR | 0 | + |
| Retucci et al <sup>33</sup> | 26 | Single Centre,<br>Milan, Italy | Respiratory<br>HDU | NIV | 29 (1 hr) | 60 | + | + | + | + | + | NR |
| Sartini et al <sup>32</sup> | 15 | Single Centre,<br>Milan, Italy | ICU/medical<br>ward | NIV | 1-3 (1-6hrs) | 60 | + | + | + | + | + | + |
| Thompson et al<br><sup>31</sup> | 29 | Single Centre,<br>NY, USA | HDU | NRB and NC | 1 hr | 60 | + | NR | + | + | + | NR |
| Tu et al <sup>30</sup> | 9 | Single Centre,<br>Shanghai, China | ICU | HFNC and<br>NIV | 3- 8 (1-4<br>hrs) | 120 | + | NR | + | + | +/- | NR |
| Xu et al <sup>29</sup> | 10 | Single Centre,<br>Anhui, China. | ICU | HFNC | 3 (16□hrs) | 300 | + | NR | + | 0 | 0 | + |
| Zang et al <sup>9</sup> | 23 | Single Centre,<br>Beijing, China | ICU | HFNC | 13.43 (8.04)<br>hrs | 30 | D | + | + | + | + | NR |
| n* - number of awake prone positioned patients in the study<br>^ PP in 1 of the 10 patients happened in medical ward following ICU consultation and supervision<br>NRB – Non-rebreather mask, hrs – hours; NC – Nasal cannula, HFNC – High-flow nasal cannula, VM – Venturi Mask / Hudson Mask, NIV – Non-invasive ventilation, ED – emergency department, ICU – intensive care unit, HDU – high dependency unit, PP – prone positioning, D – the parameter was derived from other reported values, NR – the parameter was not reported in the study, + - the parameter was reported, 0 – no events. |  |  |  |  |  |  |  |  |  |  |  |  |

In the 420 patients for whom data on the location of provision of PP was available, 111 patients (26.4%) received PP in ICU and 309 (73.6%) outside ICU (respiratory wards, high dependency units or emergency departments).

### Primary outcome

The improvements in physiological parameters (PaO_2_/FiO_2_, PaO_2_, SpO_2_) before and after PP are presented graphically in Figure 2.

**Figure 2:**
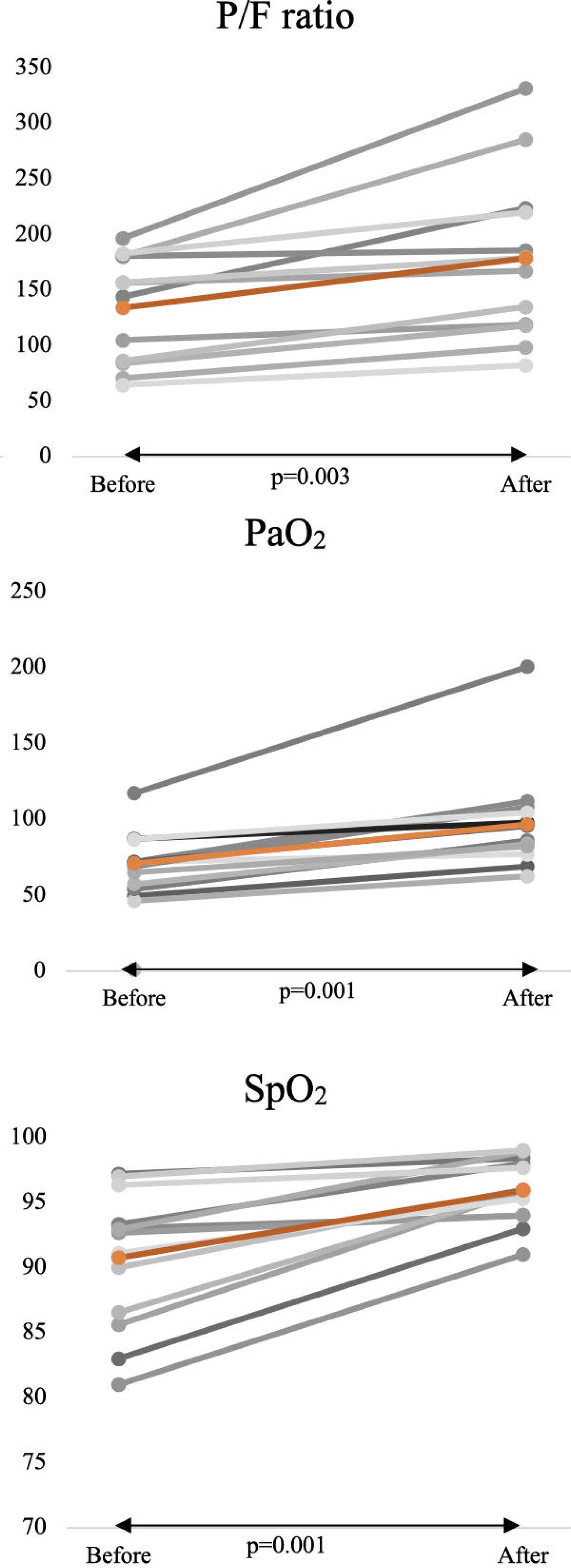
Graphical representation of mean improvements in physiological parameters post-PP

#### PaO_2_/FiO_2_ post-PP

The ratio was reported in 11 studies.^2 9 10 27-30 32 33 35 37^ The PaO_2_/FiO_2_ improved post PP (MD 37.6, 95%-CI 18.8, 56.5; p=0.001) (Figure 3). Heterogeneity persisted despite analysing studies with a sample size of more than 20 patients (4 studies,^2 9 28 33^ I^2^=97.1% p=0.001) (Supplementary Figure 1). However, the Egger’s regression test ruled out publication bias (p=0.38).

**Figure 3:**
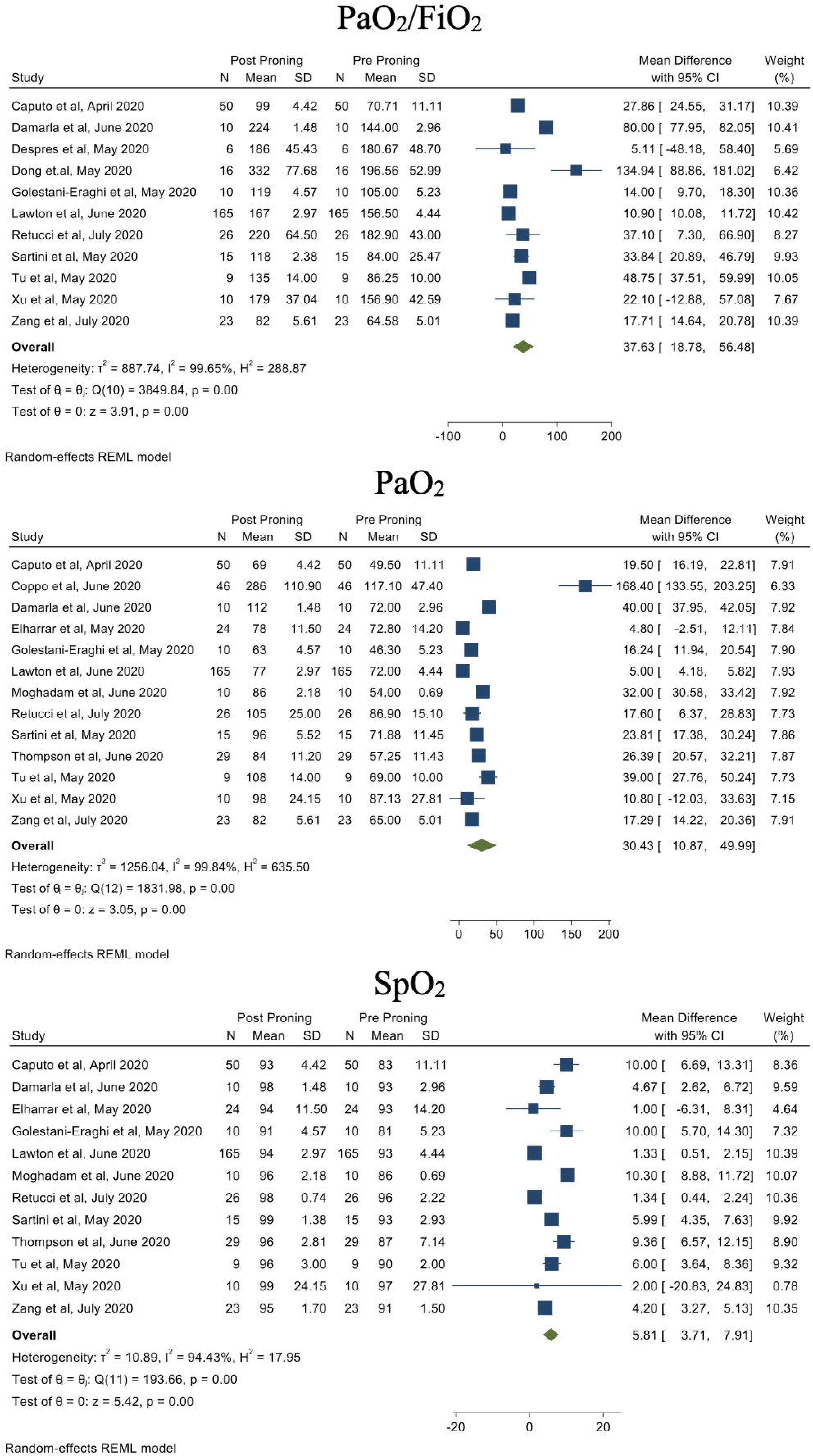
Primary Outcome demonstrating the physiological parameters post-PP

#### PaO_2_ post-PP

PaO_2_ was reported or derived from SpO_2_ in 13 studies^2 9 26-36^ (Figure 3). An improvement in PaO_2_ was demonstrated following PP (MD 30.4, 95%-CI 10.9,49.9). The heterogeneity was high (I^2^=99.8%) (Supplementary Figure 2). Egger’s regression test (p<0.001) suggests, presence of a publication bias. The heterogeneity continued to be high when only studies with more than 20 patients^2 9 26 28 31 33 36^ (I^2^=99.9%; p=0.001) were analysed.

#### SpO_2_ post-PP

SpO_2_ was reported in 12 studies.^2 9 27-36^ Improvement in SpO_2_ (MD 5.8, 95%-CI 3.7, 7.9; p=0.001) was seen across all studies where SpO_2_ was obtained (Figure 3). However, there was high heterogeneity (I^2^=94.4%) and Egger’s regression test ruled out publication bias (p=0.82). The heterogeneity continued to be high when only studies with more than 20 patients (6 studies^2 9 28 31 33 36^ I^2^=99.9%; p=0.001). (Supplementary Figure 3).

Funnel plots and Egger’s Regression test for PaO_2_/FiO_2_, PaO_2_ and SpO_2_ are presented in Supplementary Figure 4.

### Secondary Outcomes

Intubation after a trial of PP was reported in 14 studies.^2 10 26-37^ A total of 90 patients out of 426 (21.1%) were intubated following a trial of PP. The studies demonstrated moderate heterogeneity (I^2^=74.3%). The Forest plot and Funnel plot for intubation is presented in Figure 4. However, there was no publication bias (Egger’s regression test p=0.52).

**Figure 4:**
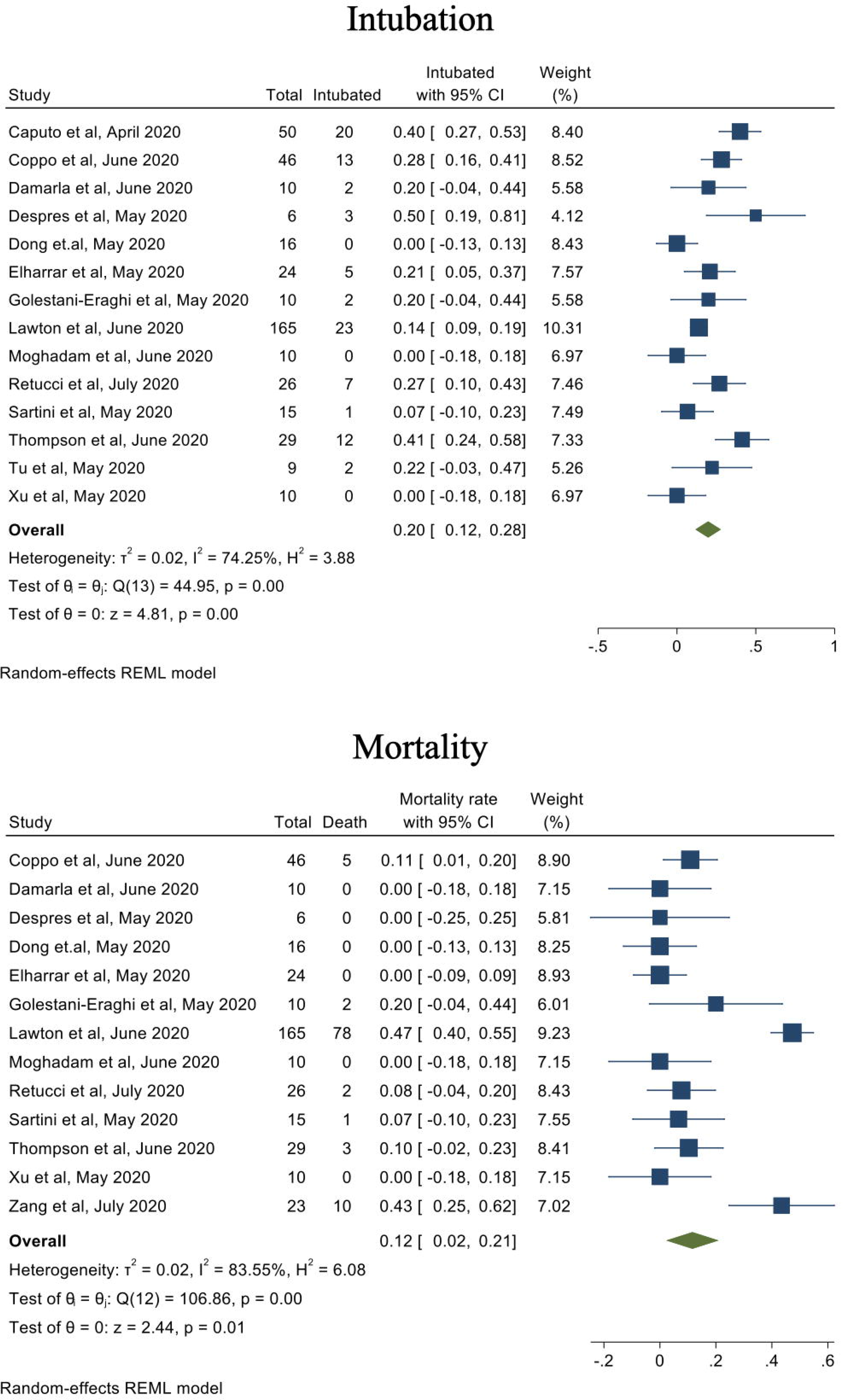
Secondary Outcomes: Forest plots for rates of intubation and mortality in patients who underwent PP.

Mortality in patients who underwent awake PP was reported in 13 studies.^9 10 26-29 31-37^ Overall, 101 patients out of 390 (25.9%) died. The studies demonstrated high heterogeneity (I^2^=83.6%), however, there was minimal publication bias (Egger’s regression test p=0.51). The Forest plot and Funnel plot for intubation is presented in Figure 4.

Funnel plots and Egger’s Regression test for intubation and mortality are illustrated in Supplementary Figure 5.

There were no reported life-threatening or major adverse events following PP. Only reported minor events included pain in the back, sternum or scrotum; general discomfort, dyspnoea and coughing and confusion in a small number of patients.^26 36 37^

Oxygenation outcomes were analysed based on the mean pre-PP PaO_2_/FiO_2_ ≤150 (5 studies^10 28 29 33 37^) or >150 (6 studies^2 9 27 30 32 35^). Patients with a Pre-PP PaO_2_/FiO_2_ ≤150 had statistically significant oxygenation improvements post-PP (MD=37 [95%-CI 17.1-56.9] vs. MD=40.5 [95%-CI −3.5-84.6]) when compared with those with a pre-PP PaO_2_/FiO_2_ >150 (Figure 5).

**Figure 5:**
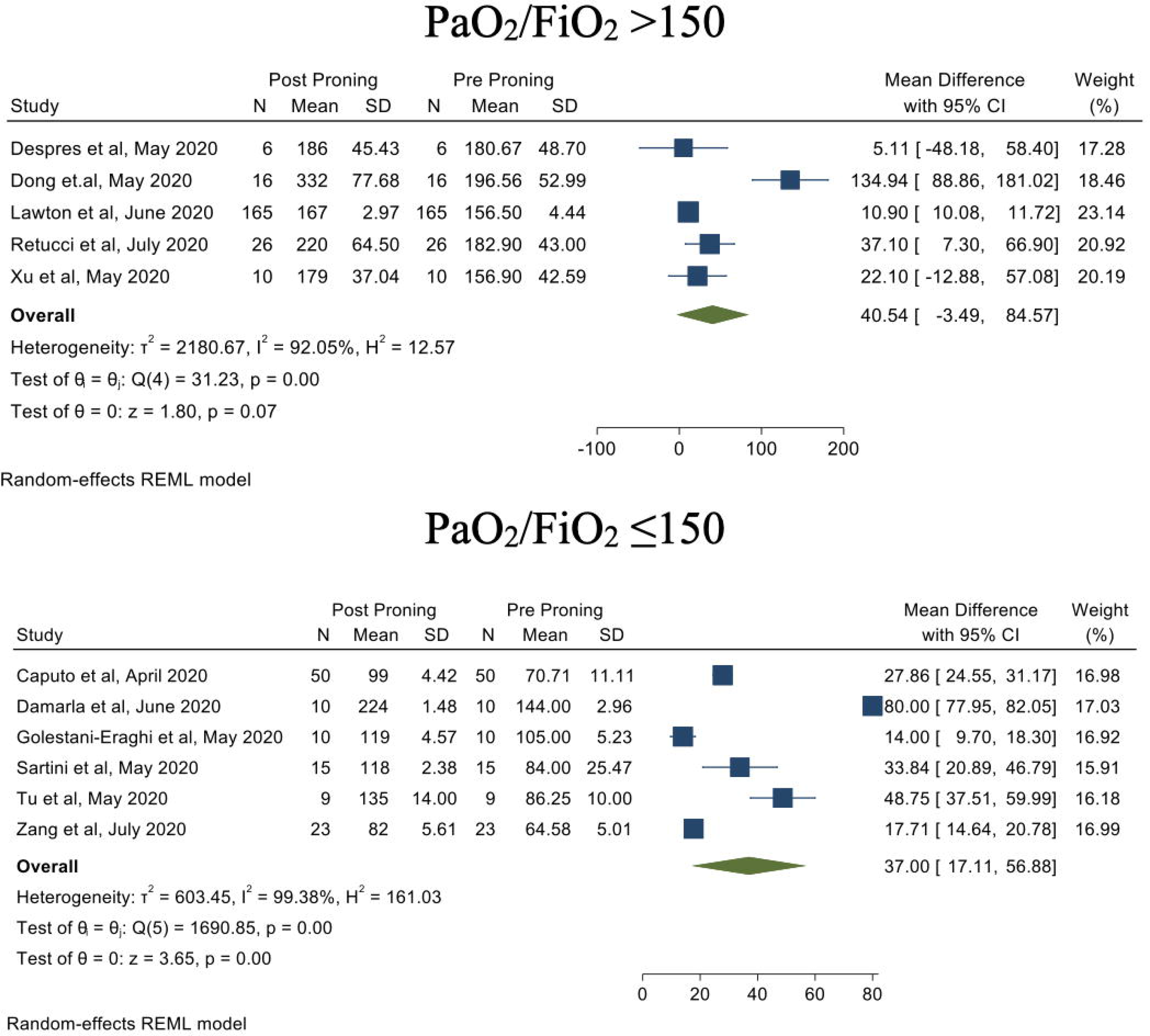
Post hoc analysis based on P/F ratio demonstrate that PaO_2_/FiO_2_ ≤150 pre-PP had statistically significant improvements when compared with PaO_2_/FiO_2_ >150

Eight studies^2 9 26-36^ reported changes in RR upon PP. There was a significant reduction in RR post-PP (MD −2.9, 95%-CI −5.4 to −0.4). High heterogenicity was observed (I^2^=93.4%) (Supplementary Figure 6) which persisted despite exclusion of smaller studies (I^2^=77.5%; p=0.01).

About a quarter of patients (111/410) received PP in ICU while others (309/410) received it in HDU, general wards and respiratory unit areas of the hospital. Physiological and clinically relevant outcomes were compared between these two locations (Supplementary Figure 7). In studies that reported on PaO_2_/FiO_2_ ratio, there was relatively higher improvement in PaO_2_/FiO_2_ in ICU patients (ICU MD=43.5 [95%-CI 11.5-75.4; p=0.001]) when compared with non-ICU patients (MD=40.8 [95%-CI 20.6–60.9; p=0.001]). PaO_2_ improvement was statistically significant in ICU patients (MD=23.8 95%-CI 14.7-32.9), whereas the improvement was insignificant in non-ICU group (MD=49.4 95%-CI −6.6-105.5). The overall improvement in SpO_2_ was 6.0% (95%-CI 3.8-8.2), however the difference was statistically insignificant between ICU (MD=5.82 95%-CI 2.46-9.16) and non-ICU (MD=6.54 95%-CI 4.31-8.76) location for PP (p=0.73). Of the 90 patients who were subsequently intubated; 64 patients (71.1%) received PP outside ICU (28.9% [26/90] in ICU vs. 71.1% [64/90]; p=0.002). Mortality data were available in 12 studies^9 10 26-29 31-34 36 37^ where patients had PP either in ICU or outside ICU. A total of 23/255 patients died (12.6% [14/111] in ICU vs. 9.6%. [9/94] in Non-ICU areas; p=0.49).

## DISCUSSION

This systematic review examined the effect of PP of non-intubated patients on oxygenation parameters in a heterogenous group of adult patients with COVID-19-related hypoxaemic respiratory failure. There was a significant improvement in oxygenation parameters (PaO_2_/FiO_2_, PaO_2_ and SpO_2_) and respiratory rate upon PP. An improvement in these parameters was consistent, although there was significant variability in both treatment dose and effect. However, due to the inconsistency of reporting physiologic outcomes, it was unclear which of these parameters may provide the best clinical guidance in terms of both patient selection for PP and evaluation of treatment response. Other relevant data for example, relative changes in patients respiratory drive, dyspnoea scores and patient comfort were not consistently available. Given these limitations, the population that clearly stands to benefit from PP could not be clearly defined.

Although all patients demonstrated improved oxygenation, the patients with PaO_2_/FiO_2_ ratio of ≤150 demonstrated a greater improvement. The reasons may possibly be that patients with more severe hypoxaemia had a greater degree of pulmonary vascular dysregulation and ventilation: those with perfusion mismatch to start with and benefited more with PP. However, such an interpretation is speculative and not much inference can be drawn from these data as an improved oxygenation with PP depends on several factors such as timing, duration, underlying pathophysiology and other respiratory supports used. For example, the duration of and frequency of prone ventilation were quite variable with some studies reporting a combination of lateral positioning and PP. Such variability is a concern when it comes to feasibility and generalisability of PP outside of centres that have some experience in PP of awake patients.

In addition, there was significant heterogeneity in oxygen therapies provided prior to and during PP. For example, 69% of the patients were receiving NIV and 12.6% were receiving oxygen via nasal cannula. These two populations can be drastically different and may represent different stages of disease evolution. This is likely to have a significant bearing on adjunctive use of PP as essentially the outcomes depend on the success of combinations of these therapies. It should be noted that ARDS studies only tested PP in intubated patients enrolled patients with a PaO_2_/FiO_2_ ≤150 to bring in some homogeneity in an otherwise heterogenous population of ARDS. In a recent network meta-analysis of trials of adult patients with acute hypoxaemic respiratory failure,^38^ treatment with non-invasive oxygenation strategies compared with standard oxygen therapy was associated with lower risk of death. Most of the included studies predated the RECOVERY trial^39^ and there was no consistent reporting on use of steroids or other disease modifying therapies limiting interpretation of the findings of the review.

Although there were no reported major adverse events following PP, not all included studies reported adverse events. Therefore, safety and efficacy of this intervention can only be tested in a well-designed randomised controlled trial and they are ongoing.^40 41^ Placing critically ill, hypoxaemic, non-intubated patients in a prone position outside closely monitored units without ability to administer invasive mechanical ventilation when required may lead to poor outcomes. PP should be carefully undertaken in systems where this can be safely provided pending further evidence. Equally, PP may be considered as a useful adjunct in patients who are considered not suitable candidates for invasive mechanical ventilation while making sure their comfort and dignity is also prioritised.

In a selected group of patients who received PP, the incidence of intubation and mortality was relatively lower in comparison with a recent systematic review and meta-analysis on associations of non-invasive oxygenation strategies and all-cause mortality in COVID-19, which reported rates of 40% and 30% respectively.^38^ In the absence of appropriate controls who did not receive PP for comparison, it is unclear whether these physiologic improvements resulted in reduced need for intubation or mortality. A noticeable difference was observed between the patients who had PP in ICU compared with other areas of the hospital both in terms of improvement in oxygenation and intubation rates. The oxygenation improvements were more marked in patients who underwent PP in ICU and there were corresponding lower intubation rates in ICU patients. However, a recent cohort study did not show any reduction in intubation rates or 28-day mortality in COVID-19 patients who received awake PP as an adjunctive therapy to HFNO.^42^ It is possible that a selected patient population of non-intubated patients with COVID-19-related respiratory failure may benefit from PP. However, data available for this review was not of sufficient quality to identify the precise population that may benefit. Based on this review, PP appears feasible and safe in patients who are hypoxaemic and when undertaken in appropriately monitored environments.

Our study has some important limitations. This review was based on data from single arm observational case series and cohort studies that had no comparator groups. Consequently, heterogeneity and all the antecedent biases associated with patient selection and reporting was expected. The heterogeneity persisted despite sensitivity analyses were performed based on sample size. Given the inconsistent reporting of oxygenation parameters, we had to derive some of the variables from other reported variables and where possible requested missing data from the corresponding authors of the included studies. Despite this, we still had missing variables in some of the included studies. This calls for a validated system to report changes in physiologic parameters in future studies that test respiratory supports in non-intubated patients. Healthcare worker infection risks and rates while assisting/facilitating PP were not reported in any of the studies. In addition, strong conclusions cannot be reached due to several factors: first, the absence of tested, established triggers and a standardised process for initiating PP in non-intubated COVID-19 patients; second, the significant heterogeneity in the patient populations included and lack of granular data on co-interventions used (NIV, HFNC, Steroids, Antiviral therapies etc.); third, an absence of standardised intubation criteria; and, fourth, that the intervention was provided in some instances under pandemic stressors that affected resource availability.

## CONCLUSION

There was a variable but significant improvement in oxygenation parameters with PP in non-intubated, hypoxic adult patients with COVID-19-related hypoxaemia. This review observed a lack of a standardised process for PP in non-intubated patients. Significant heterogeneity, inconsistent reporting, poor data quality and potential biases in data may affect the analysis. Absence of standardised intubation criteria and the provision of the intervention under pandemic stressors further limit interpretation. Well designed, randomised control studies testing the efficacy of PP in non-intubated COVID-19 patients are needed prior to widespread adoption of this practice.

## Data Availability

Availability of data and material:
All the data analysed in this study is included in the main article and supplementary files. Additional information that was obtained from some of the studies after contacting the authors is available from the corresponding author on reasonable request.

## Declaration

### Availability of data and material

All the data analysed in this study is included in the main article and supplementary files. Additional information that was obtained from some of the studies after contacting the authors is available from the corresponding author on reasonable request.

### Authors’ contributions

KS and AS conceived the study idea and co-ordinated the review process. MR, AS, ZL AZ and KS drafted the review protocol, conducted the systematic review, assisted with data analysis and wrote the initial draft of the manuscript. MR and AS contributed equally. SW assisted with literature search. AZ designed the summary tables. AA and BB conducted the statistical analysis and wrote sections of the manuscript. EF made significant contributions to the analysis plan. GB, RT, KR, DB and EF critically evaluated the manuscript and contributed to writing of the manuscript. All authors critically reviewed the manuscript and approved the final version prior to submission.

### Take-home message

Prone positioning in non-intubated severe COVID 19 patients demonstrated improvements in their oxygenation. However, significant heterogeneity in of duration and frequency of prone positioning and in other respiratory supports provided limit any further interpretation. Whether this improvement in oxygenation results in meaningful patient-centred outcomes needs testing in clinical trials.

## ACKNOWLEDGMENT

We thank authors of all the studies for providing us with the data needed for our systematic review and meta-analysis. We are also grateful to Dr. Ata Mahmoodpoor, Dr. Xu Q, Dr. Tom Lawton, Dr. Caputo for responding to our request for additional information used in this study. Prof Shekar acknowledges the Metro North Hospital and Health Service for research support.

---

**Table 1:** 15 studies included in this systematic review and meta-analysis.

**Figure 1:** PRISMA (Preferred reporting items for systematic reviews and meta-analyses) flowchart of study inclusions and exclusions.

**Figure 2:** Graphical representation of mean improvements in physiological parameters post-PP

**Figure 3:** Primary Outcome demonstrating the physiological parameters post-PP.

**Figure 4:** Secondary outcomes: Forest plots for rates of intubation and mortality in patients who underwent PP.

**Figure 5:** Secondary Analysis based on P/F ratio demonstrate that PaO_2_/FiO_2_ ≤150 pre-PP had statistically significant improvements when compared with PaO_2_/FiO_2_ >150.

**Supplementary Figure 1:** Sensitivity analysis to minimise heterogeneity for PaO_2_/FiO_2_ based on study sample size.

**Supplementary Figure 2:** Sensitivity analysis to minimise heterogeneity for PaO_2_ based on study sample size.

**Supplementary Figure 3:** Sensitivity analysis to minimise heterogeneity for SpO_2_ based on study sample size.

**Supplementary Figure 4:** Funnel plots and Egger’s regression tests for PaO_2_/FiO_2_, PaO_2_ and SpO_2._

**Supplementary Figure 5:** Secondary Outcomes: Funnel plots for rates of intubation and mortality in patients who underwent PP.

**Supplementary Figure 6:** Secondary Outcomes: Reduction in respiratory rates who underwent PP. Graphical representation of mean of mean difference pre and post-PP along with Forest plot, Funnel plot, Egger’s regression and sensitivity analysis for study sample size >20.

**Supplementary Figure 7:** Secondary Outcomes: Analysis of physiological parameters (P/F ratio and SpO_2_) based on patients’ location (ICU vs non-ICU areas) when PP was attempted.

**Supplementary Table 1:** Search terms and search engines used for the systematic review

**Supplementary Table 2:** Quality Assessment and risk of bias in individual studies evaluated using NOS and JBI Critical appraisal chest list.

**Supplementary Table 3:** Equation used for conversion of PaO2 to SPO2 and deriving SPO2 from PaO2.

**Supplementary Table 4:** Equation used to calculate mean and standard deviation from median and Inter quartile range.

**Supplementary Table 5:** Diversity in oxygen delivery modes and variation in FiO2 in the study participants.

**Supplementary Table 1:** Search terms and search engines used for the systematic review

|  |  |
| --- | --- |
| **Pubmed:** | "severe acute respiratory syndrome coronavirus 2"[Supplementary Concept] OR "COVID-19"[Supplementary Concept] OR "Coronavirus Infections"[MeSH Terms] OR COVID-19[Title/Abstract] OR "sars cov-2" [Title/Abstract] OR MERS-CoV[Title/Abstract] OR hCov*[Title/Abstract] OR "corona virus disease"[Title/Abstract] OR ((coronavirus*[Title/Abstract] OR ncov[Title/Abstract] OR sars-cov[Title/Abstract]) AND (2019[Title/Abstract] OR 19[Title/Abstract] OR Wuhan[Title/Abstract] OR human[Title/Abstract] OR MERS[Title/Abstract]))<br>AND<br>("Prone position"[Mesh]) OR ("prone position*"[Title/Abstract] OR proning[Title/Abstract]) |
| **Scopus:** | TITLE-ABS-KEY((COVID-19 OR "sars cov-2" OR MERS-CoV OR hCov* OR "corona virus disease" OR ((coronavirus* OR ncov OR sars-cov) W/3 (2019 OR 19 OR Wuhan OR human OR MERS)))) AND ("prone position*" OR proning)) |
| **Embase:** | ('covid 19'/exp OR 'coronavirus disease 2019'/exp OR 'coronavirus infection'/exp OR 'severe acute respiratory syndrome coronavirus 2'/exp OR 'covid 19':ab,ti OR 'sars cov-2':ab,ti OR 'mers cov':ab,ti OR hcov*:ab,ti OR 'corona virus disease':ab,ti OR (((coronavirus* OR ncov OR 'sars cov') NEAR/3 (2019 OR 19 OR wuhan OR human OR mers)):ab,ti)) AND ('prone position'/exp OR 'prone position*':ab,ti OR proning:ab,ti) |
| **Living systematic review:** | "severe acute respiratory syndrome coronavirus 2"[Supplementary Concept] OR "COVID-19"[Supplementary Concept] OR "Coronavirus Infections"[MeSH Terms] OR COVID-19[Title/Abstract] OR "sars cov-2" [Title/Abstract] OR MERS-CoV[Title/Abstract] OR hCov*[Title/Abstract] OR "corona virus disease"[Title/Abstract] OR ((coronavirus*[Title/Abstract] OR ncov[Title/Abstract] OR sars-cov[Title/Abstract]) AND (2019[Title/Abstract] OR 19[Title/Abstract] OR Wuhan[Title/Abstract] OR human[Title/Abstract] OR MERS[Title/Abstract]))<br>AND<br>("Prone position"[Mesh]) OR ("prone position*"[Title/Abstract] OR proning[Title/Abstract]) |

**Supplementary Table 2:**
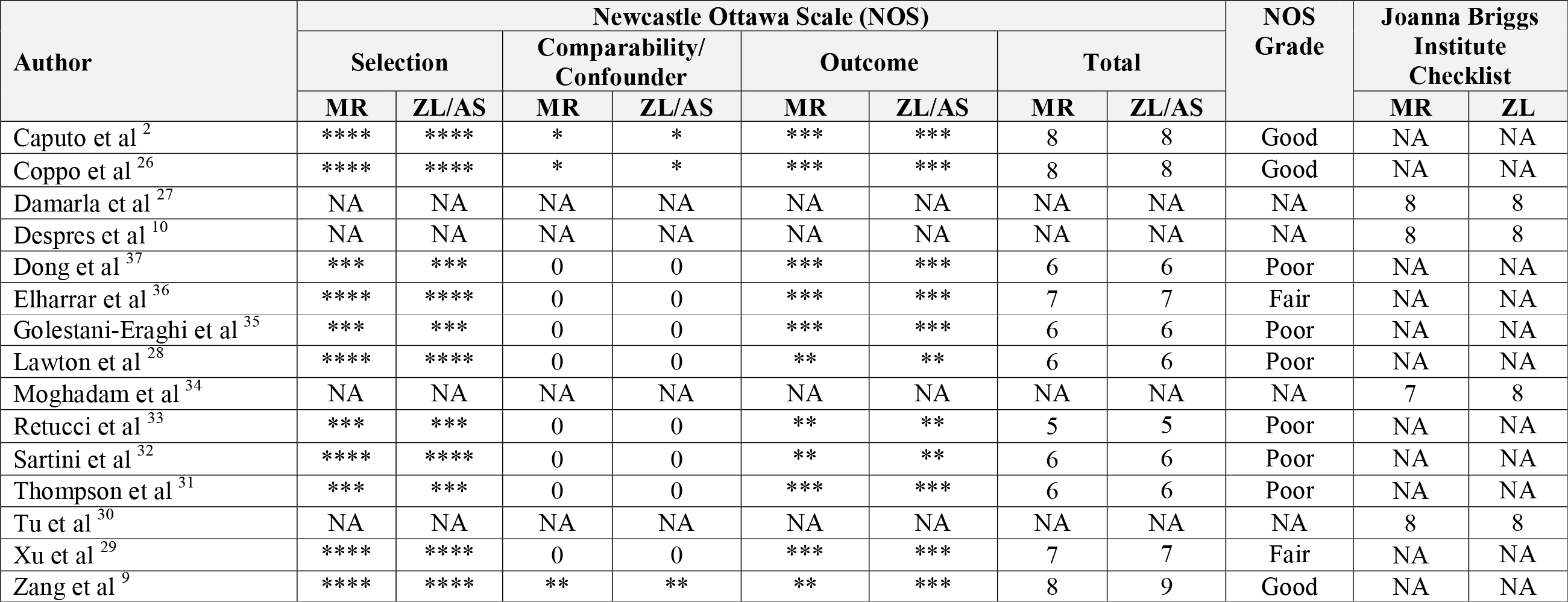
Quality Assessment and risk of bias in individual studies evaluated using Newcastle Ottawa Scale and Joanna Briggs Institute Critical appraisal checklist.

**Supplementary Table 3:** Equation used for conversion of PaO_2_ to SPO_2_ and deriving SPO_2_ from PaO_2_.

| SpO<sub>2</sub> (On monitor) | Calculation for PaO<sub>2</sub> | Resultant PaO<sub>2</sub> range |
| --- | --- | --- |
| 100% - 90% | Decrease PaO<sub>2</sub> by 4mmHg for every single percent reduction in SpO<sub>2</sub> | 100 - 60 mmHg |
| 90% - 80% | Decrease PaO<sub>2</sub> by 1.5mmHg for every single percent reduction in SpO<sub>2</sub> | 60 - 45 mmHg |
| <80% | Divide SpO<sub>2</sub> by 2 to reach to a PaO<sub>2</sub> level | 40 mmHg and downward |
| Adapted from “Correlation between the levels of SpO<sub>2</sub> and Pa O<sub>2</sub>” by A. Madan 2017 Lung India, 34(3), p.307-308 copyright by Lung India.<sup>21</sup> |  |  |

**Supplementary Table 4:**
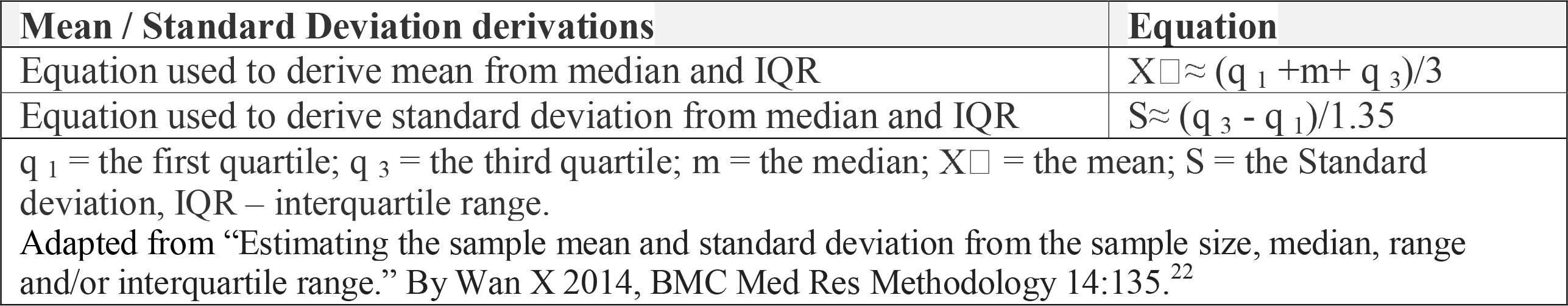
Equation used to calculate mean and standard deviation from median and Inter quartile range.

**Supplementary Table 5:** Diversity in oxygen delivery modes and variation in FiO2 in the study participants.

| Oxygen delivery mode | Reported patients (N=350) |
| --- | --- |
| NIV | 241/350 (68.9%) |
| HFNC | 17/350 (4.9%) |
| Hudson Mask | 48/350 (13.7%) |
| Nasal Cannula | 44/350 (12.6%) |
| Fraction of inhaled Oxygen (FiO<sub>2</sub>) | Reported patients (N=277) |
| >0.70 | 56/277 (20.2%) |
| 0.70-0.50 | 46/277 (16.6%) |
| <0.50 | 175/277 (63.2%) |
| NIV = Non-Invasive Ventilation, HFNP= High Flow Nasal Cannula |  |

