## Supplementary figures and images for "Prone positioning of non-intubated patients with COVID-19 - A Systematic Review and Meta-analysis"

### Supplementary Figure 1.jpeg

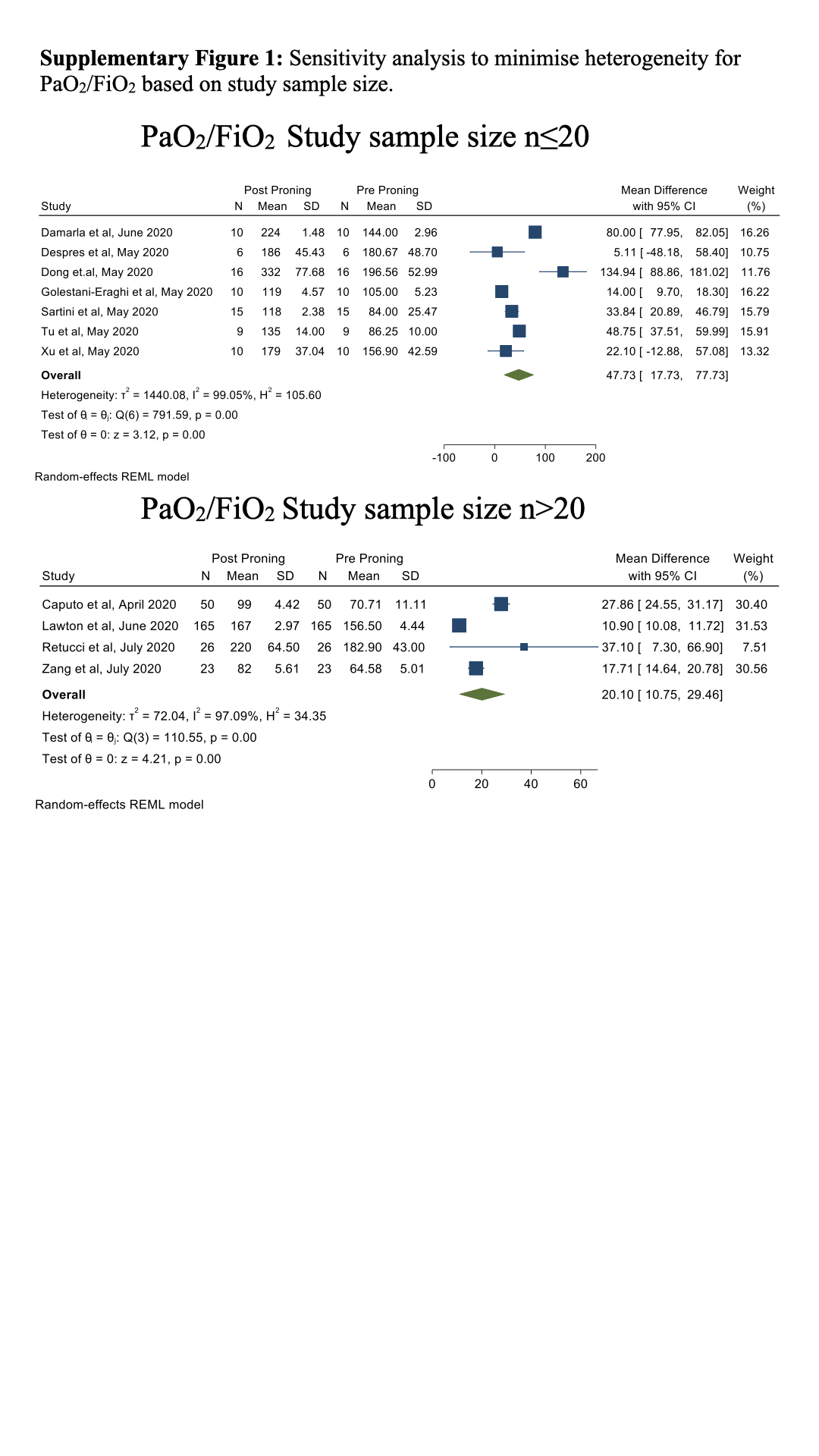

### Supplementary Figure 2.jpeg

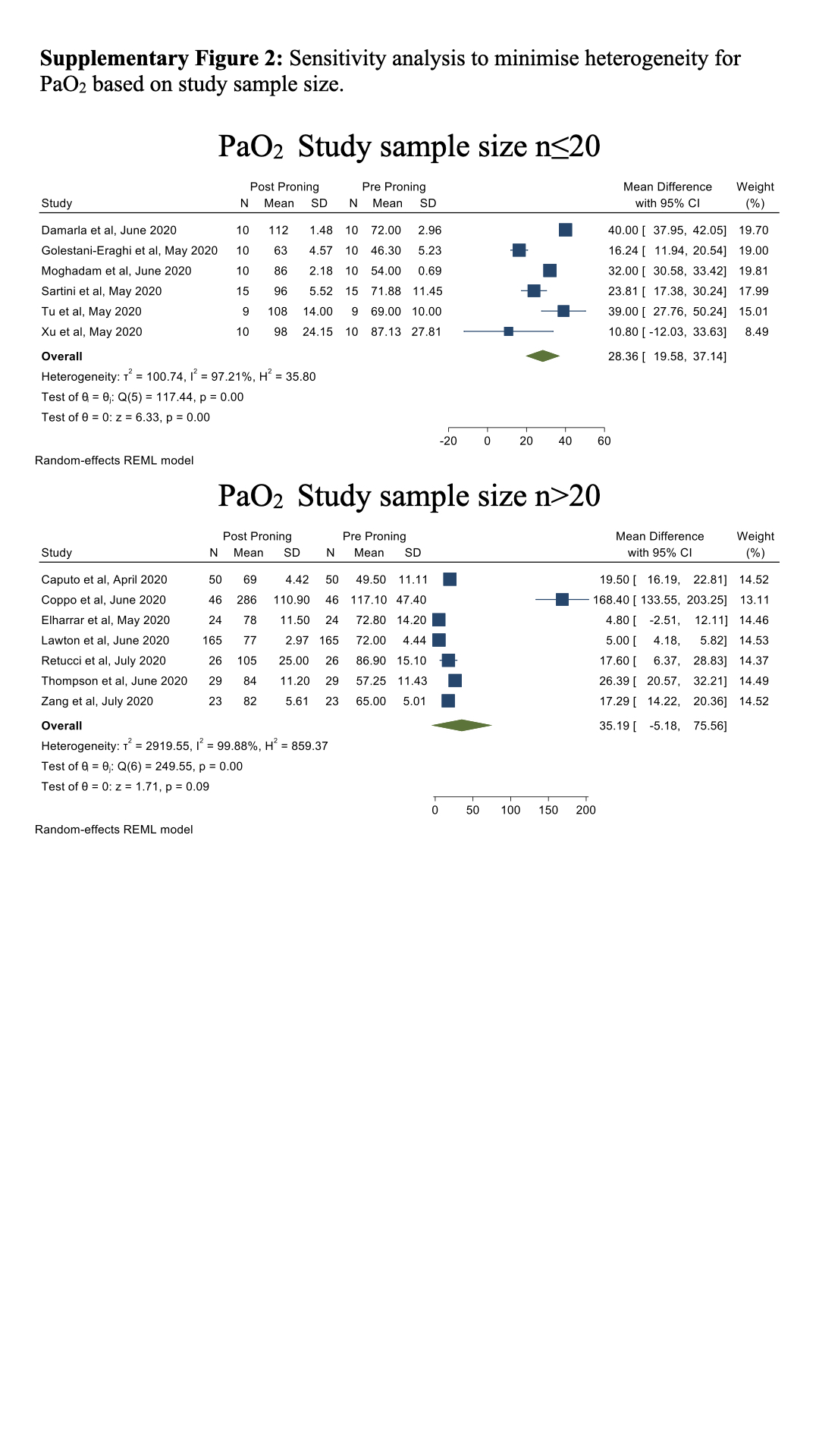

### Supplementary Figure 3.jpeg

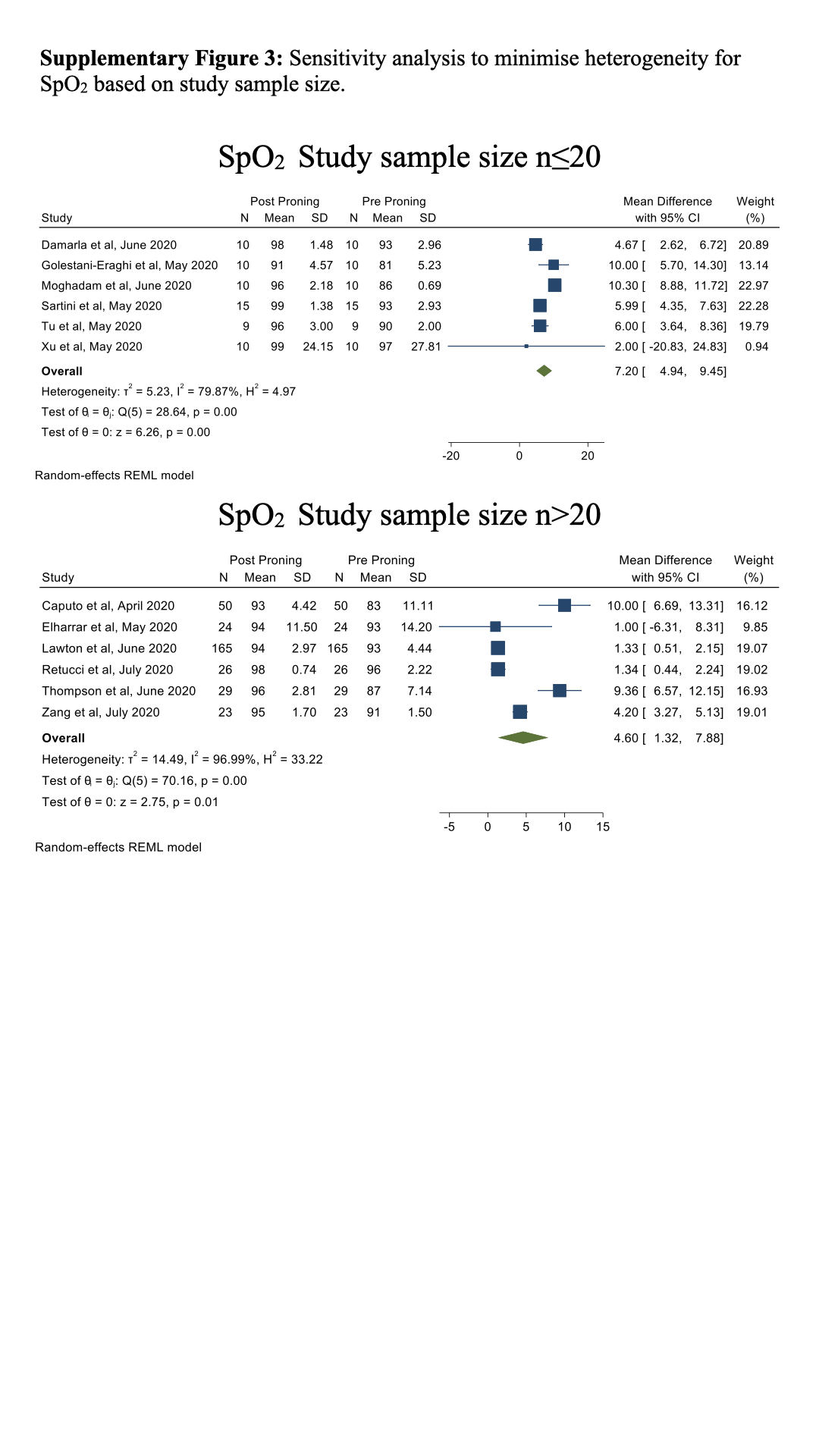

### Supplementary Figure 4.jpeg

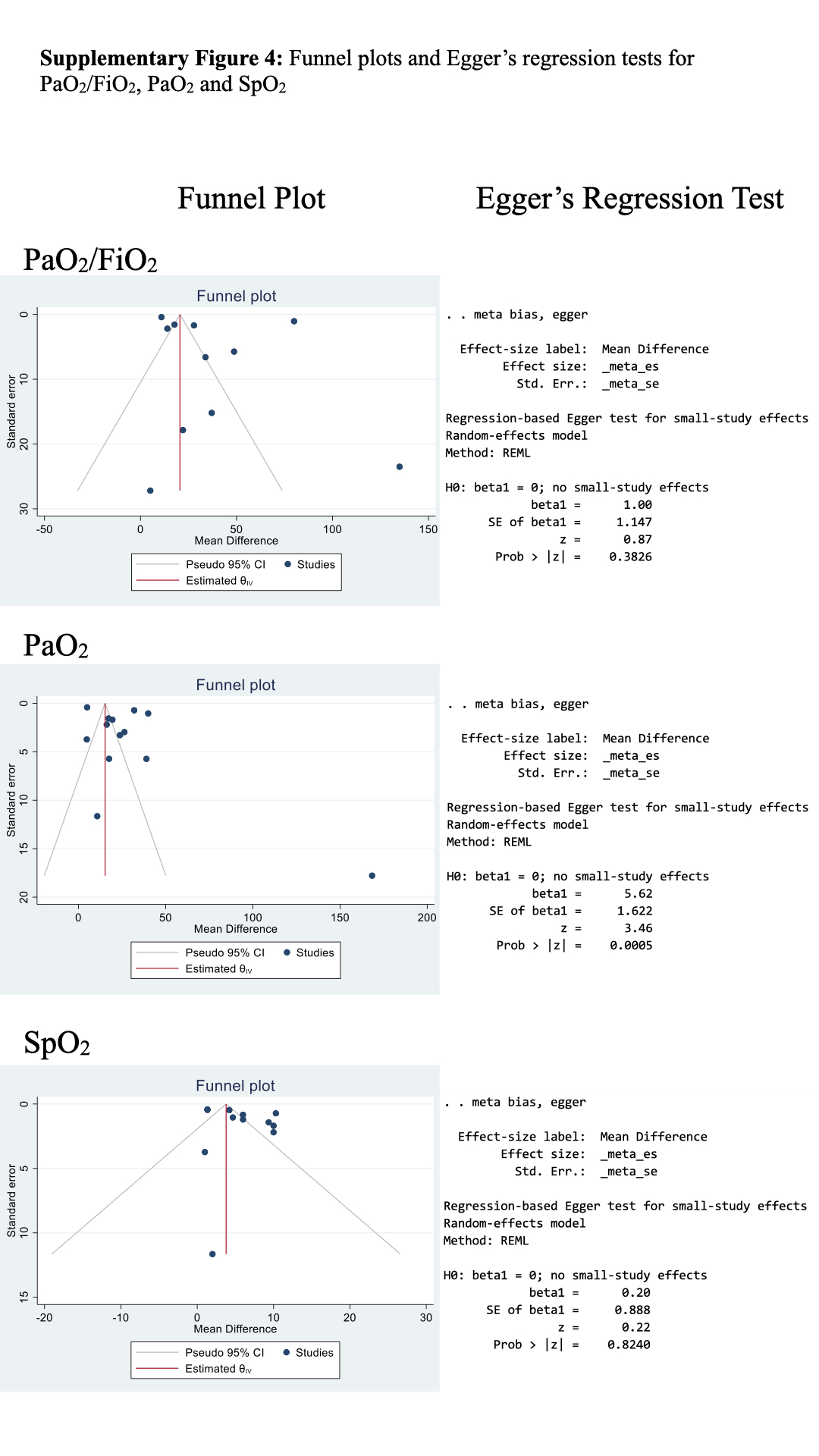

### Supplementary Figure 5.jpeg

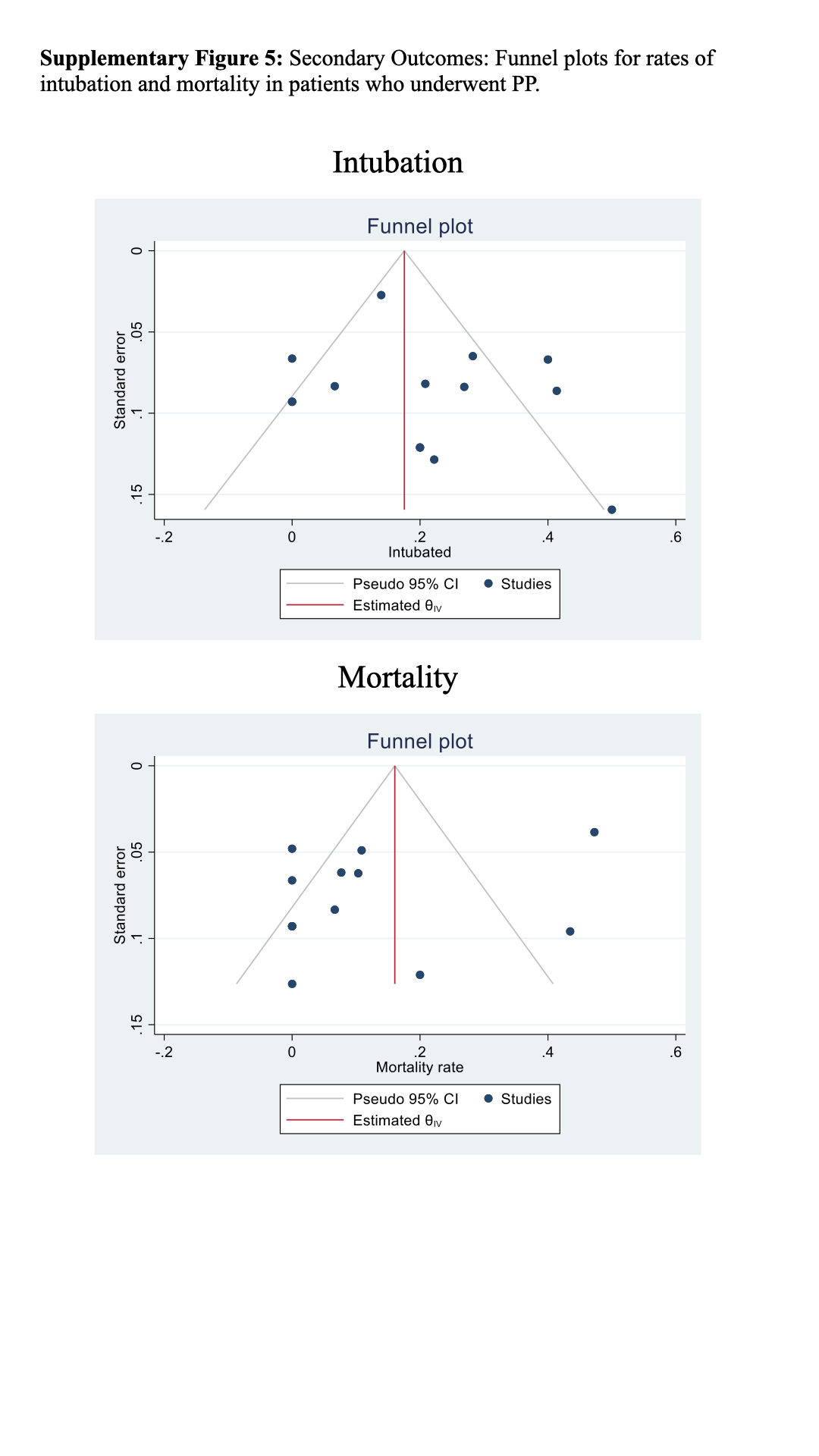

### Supplementary Figure 6.jpeg

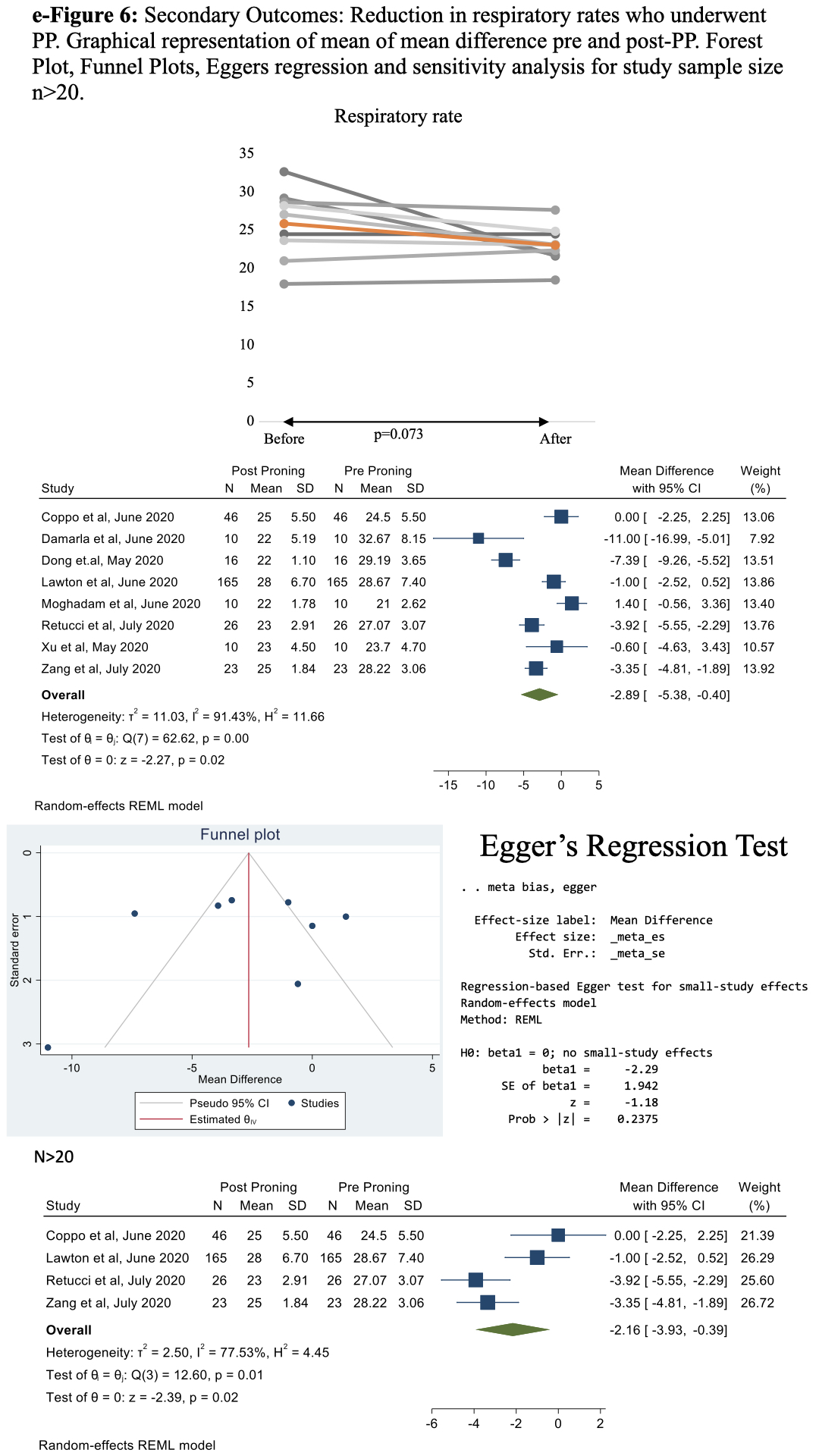

### Supplementary Figure 7.jpeg

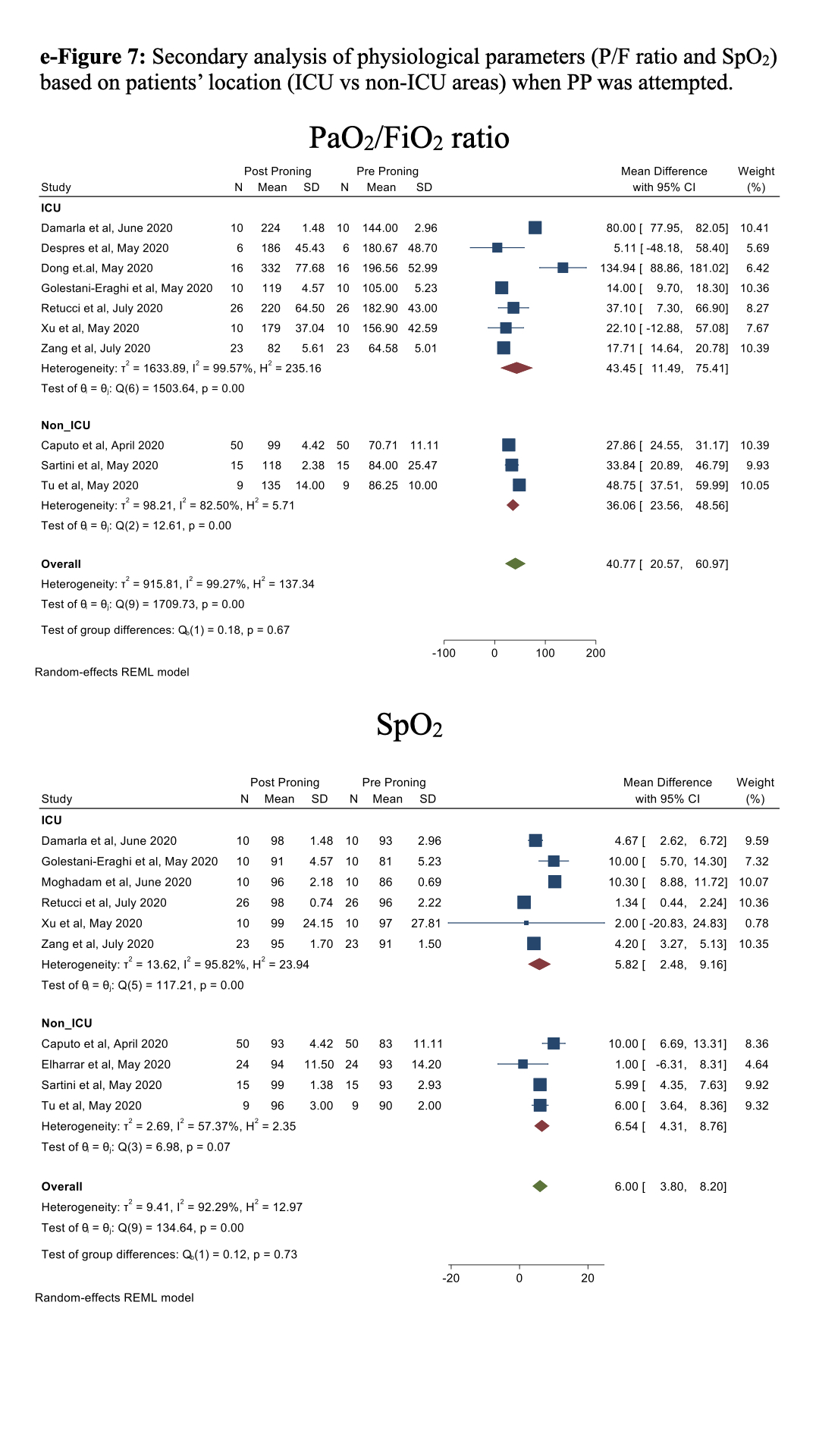
